# Self-harm incidents and restrictive practice use in an acute female inpatient unit: a service evaluation of a continuous multi-system improvement approach

**DOI:** 10.64898/2026.10.19.26365708

**Authors:** Sara Johnson, Marlene Kelbrick, Chris Griffiths, Arwa Al-Falahi, Nick Mann, Trish Ribano, Kirsty Hart

**Affiliations:** Northamptonshire Healthcare NHS Foundation Trust, Northampton, UK

## Abstract

**Background:** Self-harm is a frequent and challenging occurrence in adult inpatient psychiatric units. The use of restrictive practice to manage self-harm incidents continues to be widespread within mental health environments despite increasing national and organisational guidance to minimise its use.

**Purpose/aim:** The aim of this project was to introduce and evaluate a continuous quality improvement programme to progressively reduce incidents of self-harm and the use of restrictive practice. The project was set in a female acute mental health inpatient unit. The project sought to embed improvements in an inpatient unit’s interventions, procedures, and culture and staff’s mindset, and move towards a sustained collaborative and positive therapeutic and communication-based approach.

**Methods:** A retrospective service evaluation design was employed to assess differences in the number of incidents of self-harm, enhanced observation, seclusion, restraint and rapid tranquillisation in the period following the introduction of a continuous whole-systems multimodal interventional initiative in 2019. Pre- and post-initiation data were compared (2017/18 compared to 2019/2025) to evaluate improvement and whether this had been sustained. Data were analysed using descriptive statistics, including frequencies, percentages, and means.

**Results:** Self-harm incidents reduced between 2017/18 and 2019, with a downward trend sustained between 2019 and 2025 (136 to 72 incidents respectively between 2019 and 2025; 47% reduction). A sustained reduction in the use of restrictive practice was shown in all domains pre- and post-initiation (2017/18 compared to 2025).

**Clinical implications:** No one approach will successfully reduce self-harm and the use of restrictive practice. Approaches employed have to be multi-faceted, multi-interdisciplinary, collaborative, and blend together to deliver improvement. Sustained progress requires commitment to an iterative process of continuous collaboration, improvement and adaptation.

## 1. Background

Self-harm is defined as any act of self-poisoning or self-injury carried out by an individual irrespective of their motivation (NICE, 2022). Frequent incidents of self-harm are associated with alcohol use disorder, emotionally unstable personality disorder (EUPD) and depression (Colle et al., 2020; Sadath et al., 2023). Self-harm is a frequent and challenging occurrence in adult inpatient psychiatric units: up to 70% of patients will have self-harmed at least once during an inpatient admission (James et al., 2012; NHS England, 2025). Self-harm is often in response to psychological distress (James et al., 2012), and the inpatient ward environment can contribute to self-harm behaviour related to interactions with other service users, ward rules and restrictions including leave arrangements, high stimulus levels, and not feeling supported or safe (Nawaz et al., 2021). Risk of attempted or completed suicide is increased in people with previous incidents of self-harm (Nawaz et al., 2021). Self-harm is more common in females (Knipe et al., 2022). The United Kingdom (UK) has one of the highest rates of self-harm in Europe (Royal College of Psychiatrists, 2020). There has been a large rise in the number of recorded self-harm incidents for the UK’s National Health Service (NHS) inpatients in psychiatric wards from 24,848 in 2016 to 65,755 in 2024 (NHS Digital, 2025). Many NHS regionally based healthcare trusts fail to meet the majority of self-harm related standards (Sharp et al., 2025).

The Mental Health Act Code of Practice (1983) defines restrictive practice/interventions as ‘any deliberate acts on the part of other person(s) that restrict a patient’s movement, liberty and/or freedom to act independently to 1) take immediate control of a dangerous situation where there is a real possibility of harm to the person or others if no action is undertaken, and 2) end or reduce significantly the danger to the patient or others’ (Department of Health, 2015). Types of restrictive interventions include physical, mechanical, and chemical restraint, as well as seclusion. Enhanced therapeutic observation/engagement is also a restrictive intervention (National Mental Health Nurse Directors’ Forum 2025).

Physical restraint is defined as ‘any direct physical contact where the intervener’s intention is to prevent, restrict, or subdue movement of the body, or part of the body of another’; mechanical restraint is defined as ‘the enforced use of mechanical aids such as belts, cuffs and restraints to forcibly control a patient’s movement for the prime purpose of behavioural control’ (NHS Digital, 2024). Nursing engagement/observation is a core therapeutic intervention on mental health inpatient wards to help reduce risk to service users and staff. Observation levels are defined as low-level intermittent (30-60 minutes), high-level intermittent (15-30 minutes), continuous observation (1:1 staff member to person within eyesight or arm’s length) and multi-professional continuous (such as 2:1 staff to the person at arm’s length/within eyesight) (Aurelio et al., 2025; NICE, 2015). Seclusion is defined as ‘the supervised confinement and isolation of a patient, away from other patients, in an area from which the patient is prevented from leaving’ (Department of Health, 2015; NHS Digital, 2024).

Restrictive practices (including enhanced observations, seclusion, rapid tranquillisation, and physical restraint) are used in in-patient settings to manage self-harm and can cause both physical and psychological harm to patients and staff (Sharp et al., 2025). The use of restrictive practice to manage self-harm incidents continues to be widespread and common within NHS mental health environments (Dickens & Hosie, 2022; Nawaz et al., 2021). This is despite increasing national and organisational guidance and policy recommendations to reduce its use (CQC, 2023; NICE, 2022; Sharp et al., 2025).

There is increasing evidence of the negative impact of restrictive practice on service user well-being (Cusack et al., 2018). In addition, the use of restrictive practices have a negative impact on staff well-being (Lawrence et al., 2022), represented by increases in staff sickness, burnout and turnover; these factors have been correlated with increases in restrictive interventions (Chavulak et al., 2023). Thus, a reduction in the use of restrictive practices are likely to have a positive consequence on the ability to maintain staff wellbeing, reduce sick leave and burnout, and improve retention, which are important elements to support sustained efforts to reduce self-harm incidents and restrictive practice. Interventions to reduce self-harm incidents and the use of restrictive practice in managing these have included a range of therapeutic, environmental, cultural and organisational approaches, although the evidence remains lacking for recommendations for any particular approach (Nawaz et al., 2021; Reen et al., 2020; Shah et al., 2022).

Implementing comprehensive, recovery-focused self-management approaches can have positive outcomes in managing self-harm and suicidal intent (Newlove et al., 2025). For individuals with suicidality and self-harming behaviours, person-centred psychological interventions tailored to individual needs are recommended (NICE, 2022). However, only 25 % of patients in crisis care settings get offered formal psychological intervention (Newlove et al., 2025). Mixed interventions which combine individual therapy alongside ward management strategies and staff training are effective at reducing self-harm in the short and longer term (Nawaz et al., 2021). However, NHS trusts rarely define self-harm effectively; some trusts use definitions which do not reflect guidelines, and self-harm reduction remains underrepresented in policies (Sharp et al., 2025).

NICE recommends specific training for all staff who work with people who self-harm, which includes conducting psychosocial assessments, risk formulation, therapeutic observation methods, and patient engagement and rapport building, and it provides guidelines for staff supervision (NICE, 2022). Ward-based interventions (e.g. staff training) may reduce self-harm and suicide attempts (Quinlivan et al., 2025). A multidisciplinary self-harm pathway (which draws on dialectical behaviour therapy [DBT], problem-solving skills, cognitive analytical therapy, and structured clinical management [SCM]) in an acute inpatient mental health ward for adult women to help staff support patients who self-harm was found to be a useful framework for beneficial self-harm outcomes and reducing restrictive practice (Smith et al., 2023). Implementing a trusting and understanding approach helps to build a therapeutic alliance between patients and mental health professionals, which can reduce harm and risks (Royal College of Psychiatrists, 2020).

This paper describes a multi-system approach over several years as a part of continuous quality improvement to progressively reduce incidents of self-harm and the use of restrictive practice within an acute female mental health inpatient unit. The project sought to embed improvements in an inpatient unit’s interventions, procedures, culture and staff’s mindset, and move towards a sustained collaborative and positive therapeutic and communication-based approach.

## 2. Methods

The project was based in an acute female inpatient mental health unit provided by the NHS. Prior to the start of 2019, nursing interventions were based around the Safe Wards model (Bowers, 2014) and staff education through the delivery of Skills Training on Risk Management (STORM) (Gask et al. 2006). These didn’t significantly impact on improving overall quality of care related to self-harm, with no improvements in self-harm incidents and the use of restrictive practice. Prior to 2019, initial reviews to reduce blanket patient restrictions with the concept of ‘goal-focussed’ admissions were in their infancy, and initial admission psychology provision was limited (a substantial investment in 2025 significantly improved psychology provision within the wards).

In 2019, a new mental health quality initiative/strategy was implemented within a knowledge-to-action cycle framework based on a whole-systems multimodal approach, incorporating a combination of elements from evidence-based approaches (Graham et al., 2006). Following this, further interventional and improvement efforts were added year-on-year, involving a combination of environmental and therapeutic-related, clinical process, and lived experience involvement and staff training initiatives. These can be categorised into three main pillars: staff skill set enhancements and training, service user collaboration, and environmental improvements (see Figure 1). Staff training retained Safe Wards and STORM principles and complemented these with evidence-based principles of trauma-informed care (Saunders et al., 2023), compassion-focused care (Millard et al., 2023), use of DBT, and implementation of SCM (Heerebrand et al., 2021; Shin et al., 2026).

**Figure 1.**
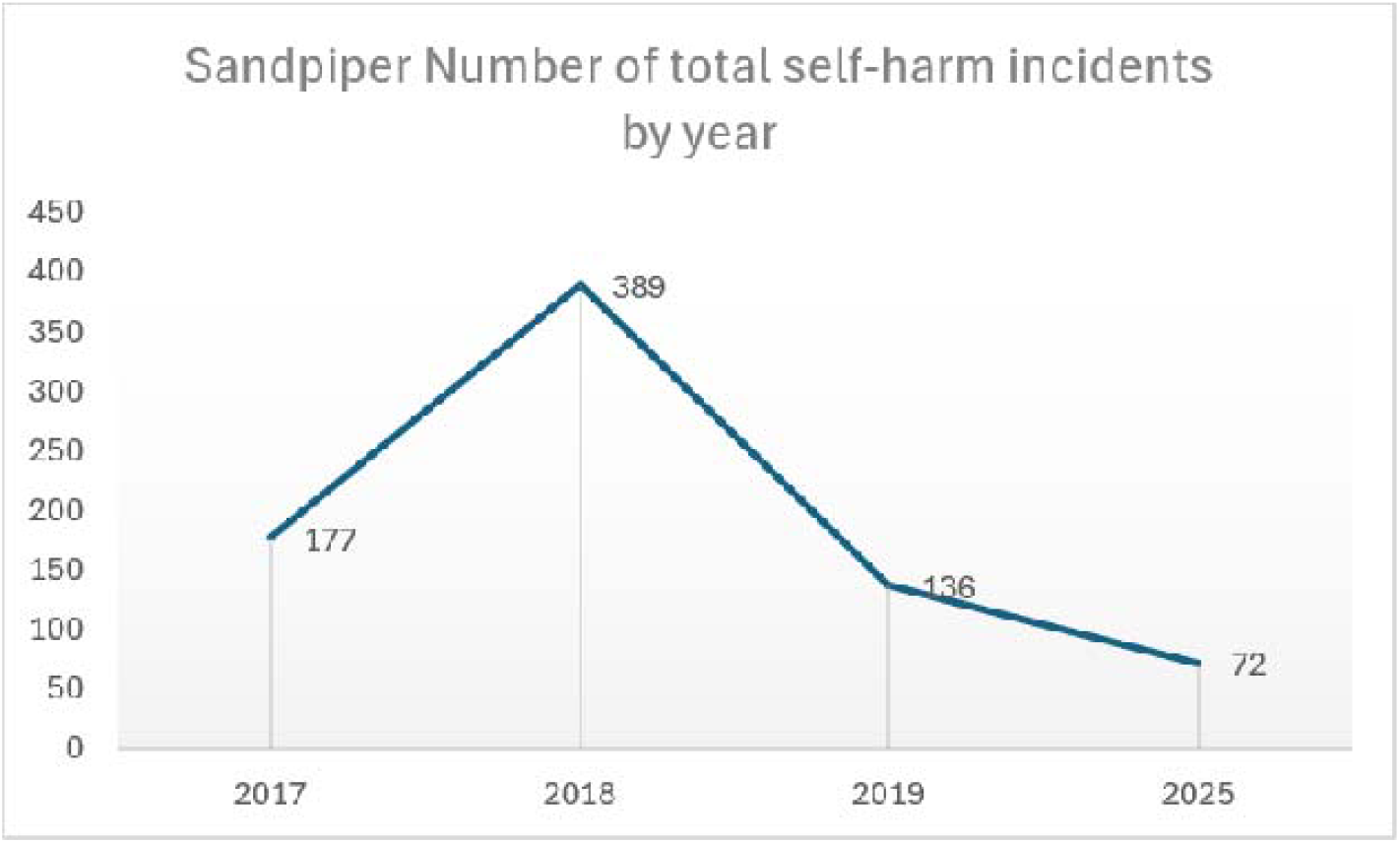

Service user collaboration was employed for implementation of interventions that promoted choice and control, positive risk-taking, and individualised safety planning. This was a move from traditional risk assessment to a keeping people safe model in line with NHS England’s mental health personalised care framework (NHS England, 2026). Service users were involved in the formulation of the function of risk behaviour and tailored responses to risk incidents. There was an introduction of weekly ward-based SCM and compassion-focused service user groups.

Environment-related improvements included ‘least restrictive’ principles adopted with individuals having access to personal belongings (no blanket restrictions but instead using an individualised approach), staff presence within communal areas, the set-up and use of a sensory modulation room, enabling individualised coping strategies, and a choice of various activity and engagement programmes. Safety pods were introduced in 2019, a safer trauma-informed alternative to physical restraint (ergonomically designed giant bean bags with head and shoulder support) (Dee’Ath et al., 2022). In terms of the structural environment, ward safety features around furniture, doors and windows were improved to reduce risk of self-harm and suicide.

An early evaluation was conducted in 2020 comparing 2017/18 and 2019 ward self-harm incidents and use of restrictive practice, with a second evaluation in 2026 comparing similar outcomes for 2025 in order to review whether improvements have been sustained. The data collected were anonymised. Data were analysed using descriptive statistics, including frequencies, percentages and means. The evaluation was agreed by the NHS Trust’s ‘Clinical Audit and Effectiveness Committee’.

## 3. Results

### Admissions

The female inpatient unit had a total of 180 admissions in 2017, 228 admissions in 2018, 222 admissions in 2019, and 138 admissions in 2025. Admissions included both direct admissions from the community and transfers from other wards. The proportion of service users with a primary diagnosis of EUPD saw an initial increase between 2017 and 2018/2019 from 19% to 27%, respectively, with a slight reduction to 25% in 2025.

### Length of stay

Length of stay (LOS) for service users with primary diagnosis of EUPD reduced between 2017, 2018 and 2019, from a mean LOS of 23 days (median 19, range 1-82 days) to a mean of 19 days (median 12, range 1-94 days) in 2018, and a mean of 12 days (median 9, range 1-79 days) in 2019. In 2025, the mean LOS in days on the ward increased to a mean of 29 days (median 21 days, range 5-136 days). This likely reflects the small number of individual patient outliers with increased complexity of mental health issues.

### Self-harm incidents

The total number of ward self-harm incidents were 177 involving 15 service users in 2017, 389 involving 39 service users in 2018, and 136 involving 10 service users in 2019. In 2025, the total number of self-harm incidents were 72, involving 16 service users (majority with primary diagnosis of EUPD: 88%). See Figure 1.

Adding 2021, 2022, 2023 and 2024 self-harm incidents, this continued to show, although fluctuating, a continuous downward trend. See Figure 2.

**Figure 2.**
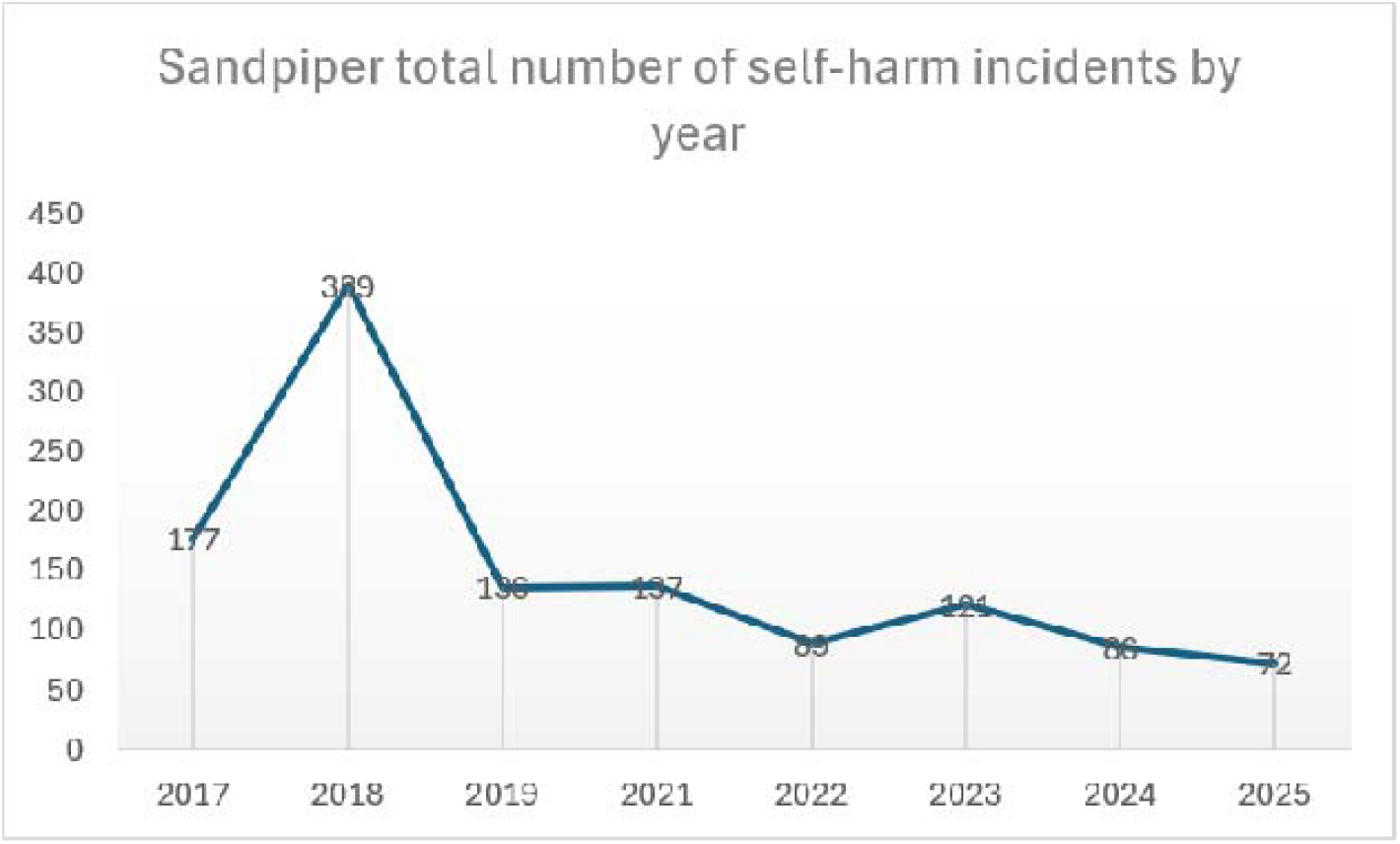

### Restrictive interventions

Enhanced ward observation/engagement use included 309 days involving 13 service users in 2017, 293 days involving 13 service users, 0 days in 2019, with 12 days (enhanced ward observation/engagement within eyesight) and 22 days (15-minute observations/engagement) involving 1 service user (different comorbid presentation) in 2025. Enhanced observation/engagement figures do not include those for safety in the context of pregnancy or physical health-related issues. See Figure 3.

**Figure 3.**
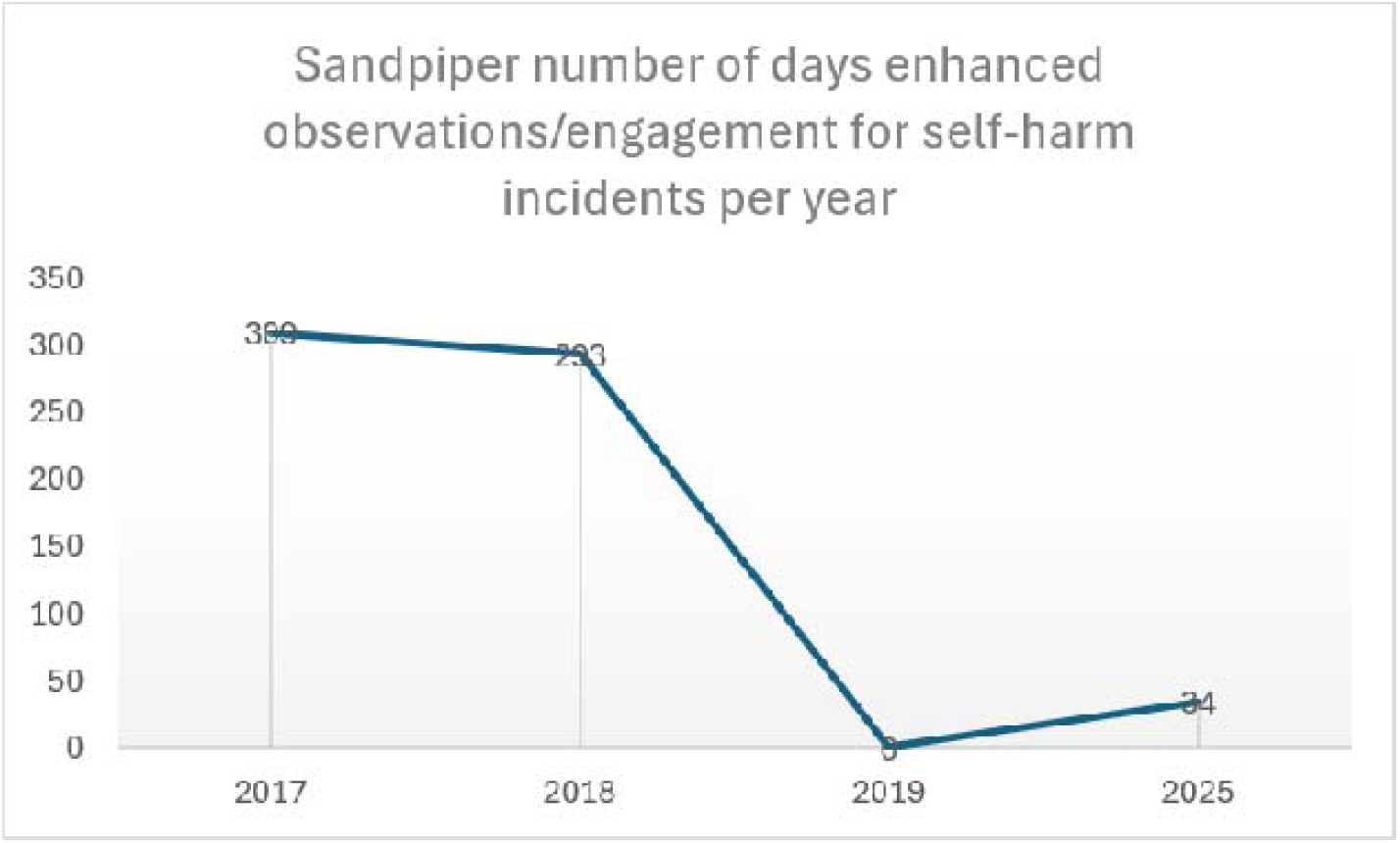

### Use of low level (LL) restraint for self-harm incidents

Low level (LL) restraint is defined as physical interaction by staff, seated or standing, such as holding of arms, usually by two members of staff, to guide or redirect a person.

There were 90 incidents of LL restraint used in 2017, 222 in 2018, 12 in 2019, and 7 incidents of use in 2025. See Figure 4.

**Figure 4.**
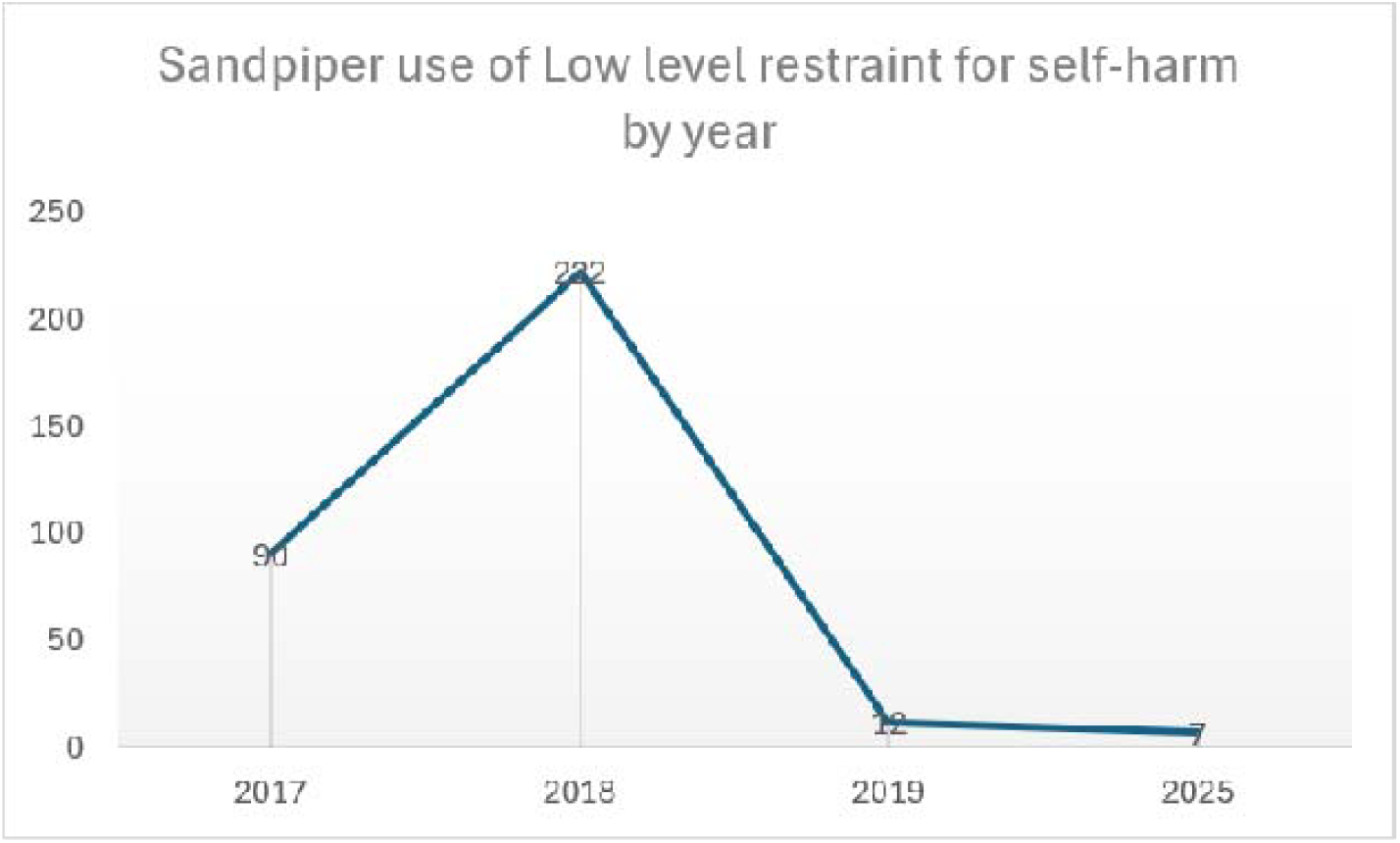

### Use of team-based restraint for self-harm incidents

Team-based restraint is defined as restraint requiring three or more trained staff to coordinate and restrict movement safely to reduce risk.

Team-based physical restraint was used in 7 incidents in 2017, 14 in 2018, and 0 incidents in 2019 and 2025. See Figure 5.

**Figure 5.**
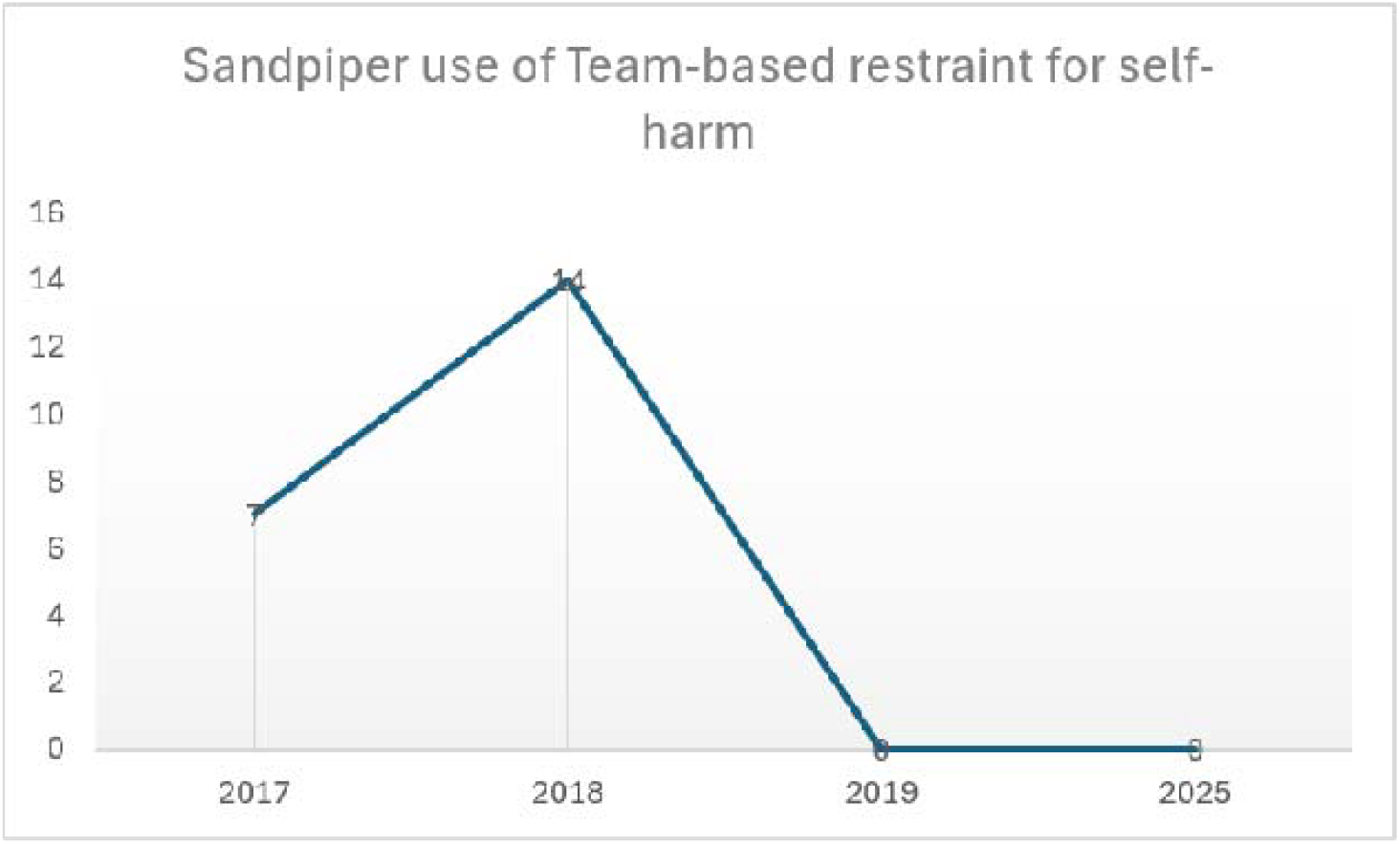

### Chemical restraint

Chemical restraint includes both PRN (“pro re nata,” or as-needed) use of medication that is defined as “the prescription and administration of medications based on the immediate patients’ needs instead of prescheduled administration times” (Mardani et al., 2022), used as part of a de-escalation or further harm prevention strategy (NICE, 2015), in this context primarily to manage acute risk behaviour. Rapid tranquillisation is defined as the ‘use of medication by the parenteral route (usually intramuscular or, exceptionally, intravenous) if oral medication is not possible or appropriate and urgent sedation with medication is needed’ (NICE, 2015).

### Use of oral PRN medication for self-harm incidents

Oral PRN medication was used in 16 incidents in 2017, 60 in 2018, 5 in 2019, and 10 in 2025. See Figure 6.

**Figure 6.**
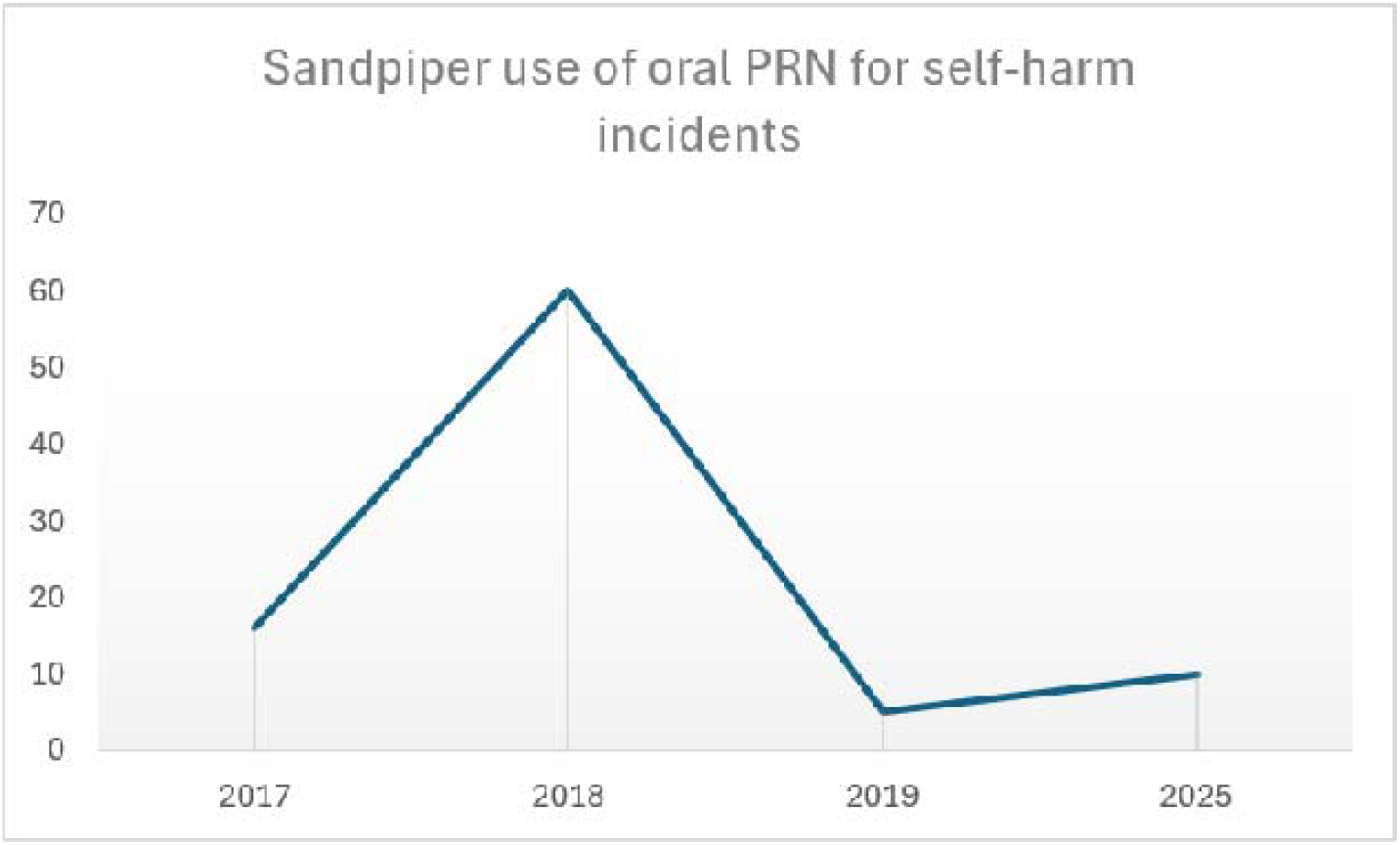

### Use of rapid tranquillisation for self-harm incidents

Rapid tranquillisation was not used for any self-harm incidents in 2017, was used for 18 incidents in 2018, and then not used for any self-harm incidents in 2019 and similarly not used in 2025. See Figure 7.

**Figure 7.**
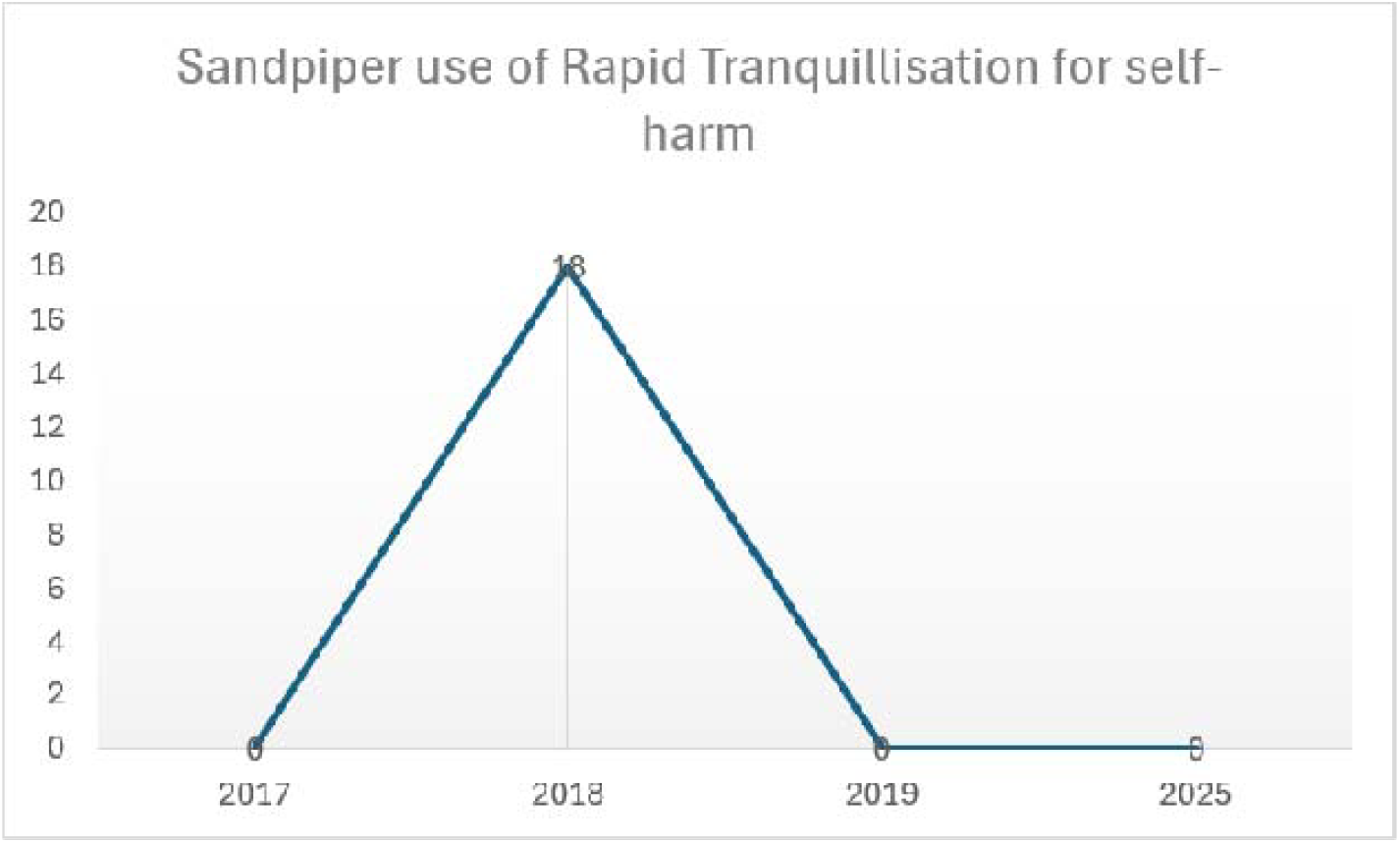

### Use of seclusion for self-harm incidents

There have been no incidents of self-harm since 2017, 2018 and 2019 that led to seclusion, and similar in 2025.

### Use of therapeutic communication emphasis approaches since 2019, evaluated in 2025

In 2025, verbal de-escalation was used in 5 self-harm incidents, the ward-based sensory modulation room was used in 12 incidents, and therapeutic 1:1 communication was used in 33 incidents.

## 4. Discussion

The project embedded improvement in an inpatient unit’s interventions, procedures, and culture and staff’s mindset, and implemented a sustained collaborative and positive therapeutic and communication-based approach. This evaluation shows an overall downward trend and sustained improvement, with reduced self-harm incidents and use of restrictive practice over a period of seven years in the female acute psychiatric inpatient unit.

The success of this continuous improvement initiative has and continues to rely on two core elements: reshaping how staff understand and respond to self-harm and embedding a cultural shift to reduce restrictive practice. The multi-layered strategy to reduce self-harm incidents and restrictive practices is built on three core pillars: proactive environmental enhancements, continuous staff development and training, and active service user collaboration. Other important factors have included workforce consistency and an effective clinical skills mix, clinical process-related factors such as embedding a Trust-wide personality disorder pathway guide and more recently implementing an initial psychological formulation for every patient on admission, with financial investment to enhance psychology provision within the inpatient units. The success of this project supports the utility and value of NICE and Royal College of Psychiatrists recommendations (NICE, 2022; Royal College of Psychiatrists, 2020). It provides evidence that supports the implementation of the findings of Quinlivan et al. (2025), Newlove et al. (2025), and Smith et al. (2023).

Meaningful service user engagement is driven by blending structured skills groups (such as SCM and compassion service user-based groups) and psychoeducation, with personalised 1:1 therapeutic communication support, and active goal setting aligned with NICE guidance (NICE, 2009; NICE, 2022). Embedding lived experience perspectives throughout all aspects of addressing self-harm can enhance relevance, acceptability and positive outcomes. Psychosocial or system-level interventions should be compassionate, patient-centred and aligned to standards (NHS England, 2024) to ensure the best outcomes.

## 5. Strengths and limitations

Northamptonshire’s general population ethnic composition is different from other UK counties, reducing generalisability. However, in terms of clinical and other characteristics, this project’s service user population is representative of a typical female acute psychiatric inpatient unit admission population. Our results are in line with other studies in terms of proportion of service users admitted to the inpatient unit with a diagnosis of personality disorder (Bredin et al., 2025; Lewis et al., 2018). Length of stay in this service user group appears similar to what has been found in other geographical areas, with the note that EUPD length of admission is influenced by individual challenging and risk behaviour, substance use, and the presence of comorbidity (Crossley & Sweeney, 2020; Flynn et al., 2024; Shoka et al., 2017; Yaniv-Rosenfeld et al., 2024 ).

## 6. Learning points

1. Reducing self-harm incidents and decreasing the use of restrictive practices are inextricably linked goals in an acute mental health inpatient setting.
2. Sustaining reduced self-harm incidents and restrictive practice use requires a multi-system continuous improvement and adjustment approach.
3. Achieving sustainable reductions in self-harm and restrictive interventions relies heavily on driving a fundamental shift in organisational culture and staff’s mindset. In this context, a paradigm shift from risk containment to autonomy-informed collaborative care.

## 7. Clinical implications

Reducing self-harm incidents and use of restrictive practice by the strategies employed is replicable in similar NHS inpatient settings. There is no single approach that will successfully reduce self-harm and the use of restrictive practice and improve patient safety. Approaches employed must be multi-faceted and multi-interdisciplinary to deliver improvement. Sustained progress requires commitment to an iterative process of continuous improvement and adaptation.

## Data Availability

Data is not available

